# A prognostic model for Senescence-related LncRNA in a novel colon adenocarcinoma based on WGCNA and LASSO regression

**DOI:** 10.1101/2025.02.13.25322203

**Authors:** Yichu Huang, Guangtao Min, Hongpeng Wang, Lei Jiang

**Affiliations:** The First Clinical Medical College of Lanzhou University, Lanzhou, China, Lanzhou, Gansu, China; General Surgery Ward 6, The First Clinical Hospital of Lanzhou University, Lanzhou, China, Lanzhou, Gansu, China

**Keywords:** colon adenocarcinoma, LASSO, WGCNA, Senescence-related LncRNA

## Abstract

**Objective:** This study aims to develop a prognostic model based on senescence-related long non-coding RNAs (lncRNAs) to predict the prognosis of patients with colon cancer and enhance their survival rates.

**Method:** Differential expression analysis and Pearson correlation were employed to identify senescence-related lncRNAs in colon cancer. A risk prognosis model was constructed using univariate Cox regression analysis and Least Absolute Shrinkage and Selection Operator (LASSO) regression analysis. The reliability of this model was validated through survival analysis, receiver operating characteristic (ROC) curves, bar charts, and calibration curves. Additionally, the relationship between the prognostic model, immune microenvironment, and drug sensitivity was explored.

**Results:** A risk prognosis model comprising eight senescence-related lncRNAs (LINC02257, AL138921.1, ATP2B1-AS1, AC005332.7, AC007728.3, AC018755.4, AL390719.3, THCAT158) was successfully established, demonstrating strong performance in predicting the overall survival rates of colon cancer patients. A significant correlation was observed between the senescence-related lncRNA progn model and the tumor microenvironment, immune infiltrating cells, and drug sensitivity.

**Conclusion:** The senescence-related lncRNA prognostic model developed in this study can accurately forecast the prognosis of colon cancer patients, offering new insights for personalized treatment approaches in colon cancer.

## 1. Introduction

Colon cancer is a type of malignant tumor influenced by various genetic and environmental factors, posing a significant threat to human health. In recent years, factors such as economic development, changes in dietary habits and lifestyles, and the rapid aging of the population have contributed to a yearly rise in both the incidence and mortality rates associated with colon cancer. The latest global cancer data in 2020 shows that[1, 2], colon cancer has the third highest incidence rate and the highest case fatality rate among malignant tumors. The symptoms of this disease often develop gradually, and the rate of endoscopic screenings relatively low in China. Most patients have lost the best surgical opportunity when seeking treatment[3, 4]. Consequently, identifying novel diagnostic and therapeutic targets for colon cancer is crucial for enhancing its diagnosis and treatment, as well as improving patient outcomes.

Aging represents the cellular response to various stress signals, serving to protect cells from unnecessary harm. In the context of cancer, aging has a dual function: it acts as a tumor suppressor by inhibiting the proliferation of damaged cells, while simultaneously promoting cancer by fostering an inflammatory environment. Furthermore, cancer cells can also exhibit aging responses. This presents both challenges and opportunities in the sequential treatment of cancer, utilizing pro-senescence therapy followed by senolytic therapy[5]. Long non-coding RNA (lncRNA) is a type of non-coding RNA that exceeds 200 nucleotides in length. It functions biologically by regulating gene expression and is crucial in the development and progression of cancer[6]. LncRNAs are instrumental in modulating various processes in colon cancer, including cell proliferation apoptosis, and cell death, as well as influencing the cell cycle migration, capabilities,esymal transition (T), cancer stem cell behavior, and resistance to colon cancer therapies[7]. E2F1-reactive lncRNA LIMp27 competes with p27 mRNA for binding to cytoplasmic hnRNP0, selectively downregulating p27 expression. This interaction results in G0/G1 phase cell cycle and promotes the proliferation, tumorigenicity, and therapeutic of colon adenarcinoma cells lacking p53[8]. Investigating senescence-related lincRNAs in colon adenocarcinoma can enhance our understanding of the molecular mechanisms involved in the onset and progression of this cancer, while also paving the way for the development of new potential intervention strategies.

## 2. Method

### 2.1 Public database acquisition and processing

We retrieved mRNA sequencing and non-coding RNA sequencing data for TCGA-COAD patients from the TCGA database, along with the clinical and pathological characteristics of colon cancer patients. Additionally, senescence-related genes associated with colon adenocarcinoma were sourced from published literature.[9].

### 2.2 Screening of Senescence-related lncRNAs

Using the R packages “limma,” “ggplot2,” “ggalluvial,” and “dplyr,” we performed Pearson correlation analysis to assess the relationship between senescence-related genes and lncRNA expression. The criteria applied were a correlation coefficient (corFilter) of 0.4 and a p-value of 0.05. A Sankey diagram was created to illustrate the correlation between senescence-related lncRNAs and senescence-related genes.

### 2.3 Construction of a prognostic model for Senescence-related lncRNA

Utilizing the R package “caret,” the adenocarcinoma cell dataset obtained from TCGA was randomly divided into a training set (n=182) and a testing set (n=273). The reliability of the prediction model was validated using the testing set and the entire TCGA dataset. To further senescence lncRNAs, single-factor Cox regression analysis was performed, and the findings were illustrated in a forest plot. Lasso regression analysis was then employed to select the optimal group of lncRNAs to develop a risk model. Subsequently, multivariate Cox regression analysis was conducted to create a prognostic model based on the selected lnc The risk score each cancer patient was calculated the equation: risk score = ∑βncRNA × Exp_lncRNA). A chi-square test was to assess whether there were statistically significant differences in clinical data between the training and testing sets. Additionally, the “ggplot2” package was used to generate a correlation heatmap depicting the relationship between senescence-related lncRNAs and senescence-related genes.

### 2.4 Validation of the efficacy of prognostic models

Patients were categorized into high-risk and low-risk groups based on the median risk score. The “survival” package was utilized to assess whether there were significant differences in overall survival (OS) between these two groups through Kaplan-Meier survival curves. To validate the independence of the model, independent prognostic analyses were conducted considering factors such as patient age, tumor grade, and stage. Utilizing the patients’ survival status along with the “surminer” and “timeROC” packages, receiver operating characteristic () curves were generated, and the area under the curve (AUC) was calculated. Additionally, a concordance index (C-index) curve was drawn to evaluate the model’s accuracy. Patients were further divided into early (stage I-II) and late (stage III-IV) stages based on tumor staging, and Kaplan-Meier survival curves were employed to compare the OS of high-risk and low-risk groups in both early and late-stage patients.

### 2.5 Column chart and principal component analysis

Column charts representing the overall survival (OS) rates for colorectal cancer patients at 1, 3, and 5 years were created using the “rms” and “sur” packages. Risk scores were integrated with clinical pathological factors for analysis. The Hosmer-Lemeshow calibration curve was employed to evaluate the predictive performance of these column charts. Additionally, the “scatterplot3d” package was utilized to conduct principal component analysis (PCA), assessing the spatial distribution of high-risk and low-risk samples based on the expression levels of all genes,escence-related genes, senescence-related lncRNAs, and lncRNAs from the risk model.

### 2.6 Evaluation of tumor microenvironment and immune cell infiltration

Tumor microenvironment analysis was conducted using the R packages “limma,” “estimate,” and “ggpubr.” Single sample gene set enrichment analysis (ssGSEA) was employed to evaluate the differences in immune cell and immune functions between high-risk and low-risk groups within the entire sample cohort. The results were visualized using “ggboxplot” package.

### 2.7 Drug sensitivity analysis

Drug sensitivity analysis is employed to identify potential drugs for the treatment of osteosarcoma (OS) patients. In this study, we utilized the R software packages “limma,” “ggpubr,” and “oncoPredict” to conduct a drug sensitivity analysis on the entire dataset, with a screening criterion of P<0.001, aiming to predict the sensitivity of potential therapeutic agents in two risk subgroups of OS patients.

### 2.8 Statistical processing

Utilizing R language (version 4), we conducted Lasso Cox regression, survival analysis, and PCA analysis. Kaplan-Meier analysis was performed with the ‘survival’ package. Furthermore, the prediction model was validated using the “survival,” “pheatmap,” and “ggpubr” packages. A P-value of less than 0.05 was considered indicative of a statistically significant difference.

## 3. Conclusion

### 3.1 Construction of a Prognostic Model for Senescence-related lncRNA Results

Randomly divide colon cancer patients into a training set (n=182) and a testing set (n=273). Table 1 shows the clinical and pathological characteristics of the patients. There was no statistically significant difference in clinical characteristics between the training and testing sets (P>0.05). Pearson correlation analysis identified 986 Senescence-related lncRNAs related to aging genes, as shown in Fig. 1A. The single factor Cox regression analysis of the training set showed a total of 9 lncRNAs strongly associated with disulfide death, and the forest plot of their risk values is shown in Fig. 1B. Lasso regression analysis and multivariate Cox regression analysis further screened out 8 lncRNAs associated with aging (Fig. 1C, D). The correlation heatmap of Senescence-related genes showed that Senescence-related lncRNAs were positively correlated with the expression of 18 genes including WNT16, IL-2, and FGF2, as shown in Fig. 1E, and negatively correlated with TUBGCP2, SCAMP4, and CD9.

**Table 1.**
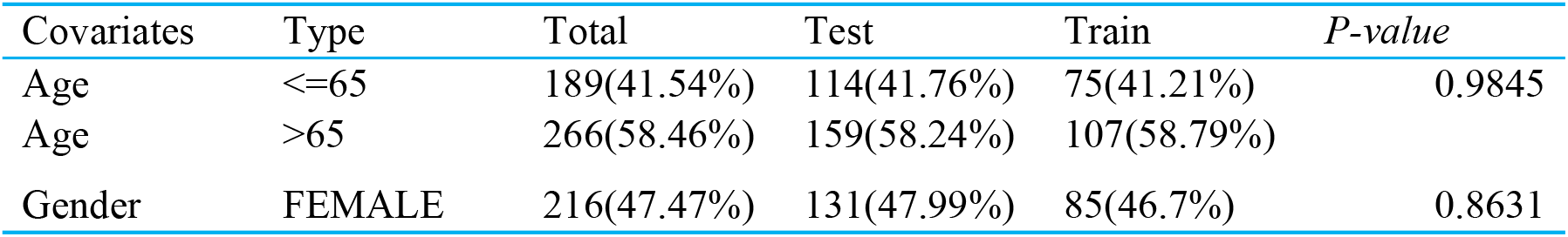
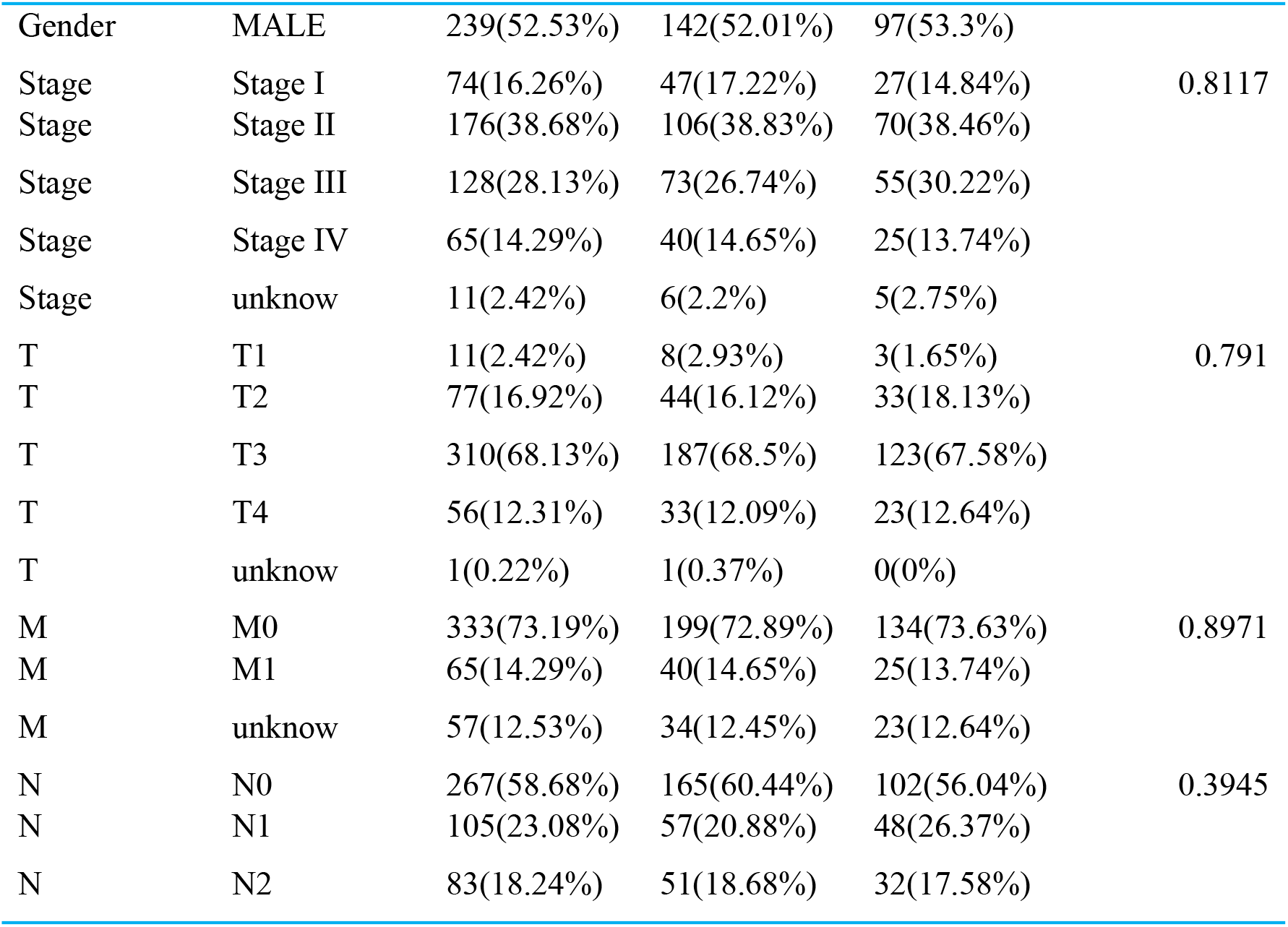
Comparison of Clinical Data between Test and Training Sets.

**Fig. 1.**
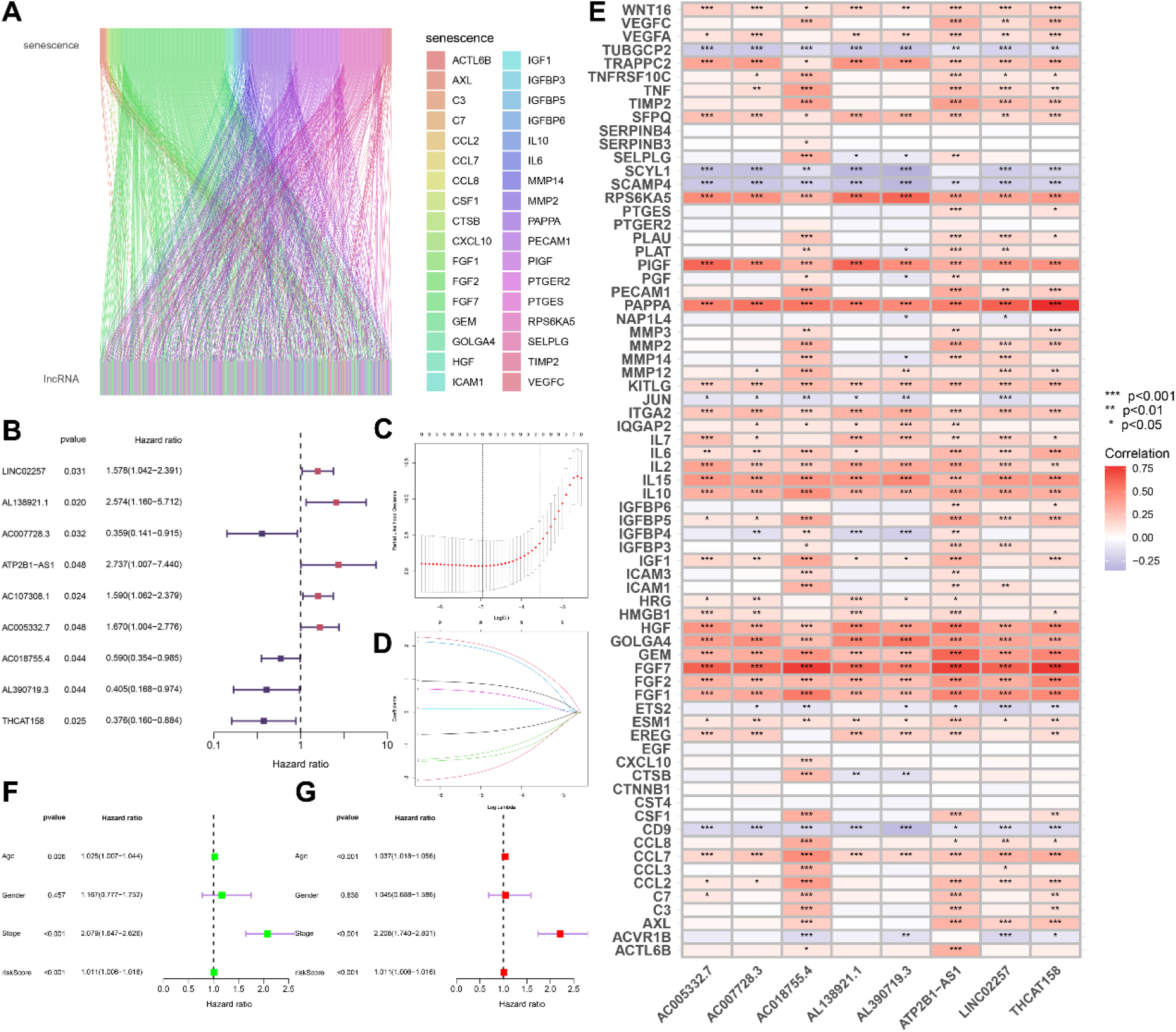
Construction of a prognostic model for senescence-associated lncRNAs A. Senescence-associated lncRNA Sankey diagram; B. One-factor Cox regression analysis; C, D Lasso regression analysis and multifactorial Cox regression analysis; E. Correlation heatmap of senescence-associated genes; F. One-factor Cox regression analysis; G. Multifactorial Cox regression analysis

### 3.2 Independent analysis of prognostic factors

To verify whether the Senescence-related lncRNA prognostic model can be used as an independent prognostic factor independent of other clinical traits, independent prognostic analysis was conducted on 8 Senescence-related lncRNAs and other clinical features involved in constructing the model. The results of univariate and multivariate Cox regression analysis showed that risk score is an independent prognostic factor compared to other clinical features (Fig. 1F, G). By combining risk score with clinical pathological features, a column chart was used to predict 1-, 3-, and 5-year survival rates, with predictive factors including risk score, age, tumor grade, and staging. The calibration chart shows that the constructed model is close to the ideal model (Fig. 2A). The C-index curve shows that the C-index index of this feature is much higher than that of Age and gender, indicating that the model has the highest accuracy in predicting patient survival (Fig. 2B, C). In addition, the ROC curve shows that the model has high accuracy in predicting patient survival rates at 1 year, 3 years, and 5 years (AUC of 0.708, 0.733, 0.771) (Fig. 3D), and its predictive ability is relatively high compared to other clinical features (AUC=0.733) (Fig. 2E). The above results indicate that the model can serve as an independent predictor and has stronger predictive ability compared to other clinical features. In order to test the predictive ability of the model for the prognosis of patients with different clinical features, survival analysis was conducted on patients with early and late stage tumors according to high and low risk. The results showed that the overall survival rate (OS) of the high-risk group was shorter than that of the low-risk group in both early and late stage patients (Fig. 2F, G). For principal component analysis of the model, based on the expression levels of all genes, Senescence-related genes, Senescence-related lncRNAs, and risk model lncRNAs, PCA analysis of the high-risk and low-risk groups showed that the risk model lncRNAs could better distinguish between the high-risk and low-risk groups (Fig. 2H-K).

**Fig. 2.**
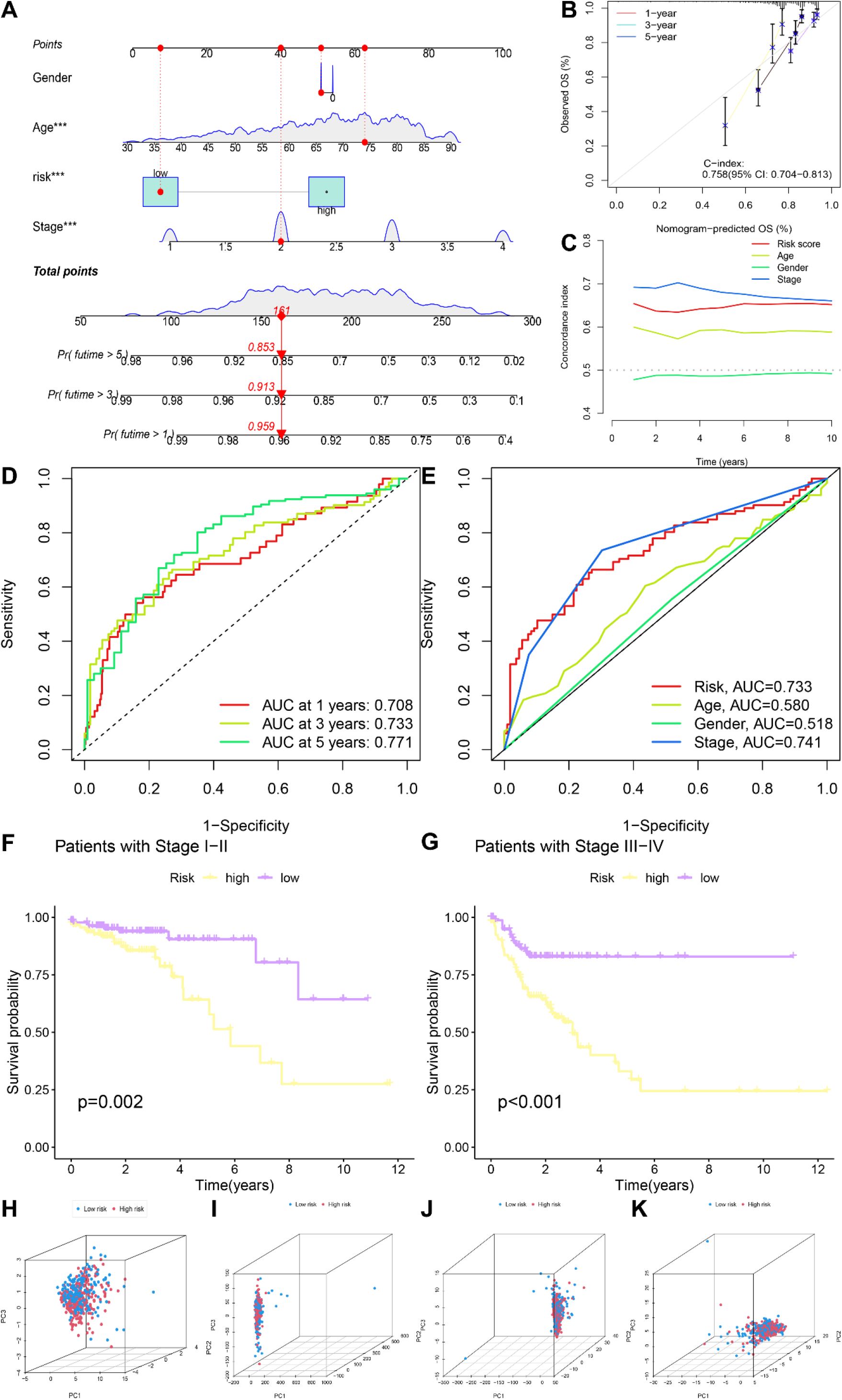
Validation of Predictive Models and Survival Analysis. A, B. Column plots; C. C-index index; D, E. ROC curves; F, G Early and Late Survival KM analyses; H Risk Model lncRNA Principal Component Analysis; Senescence-associated lncRNA Principal Component Analysis; J. Senescence-associated Gene Principal Component Analysis; K. Principal Component Analysis of All Genes.

**Fig. 3.**
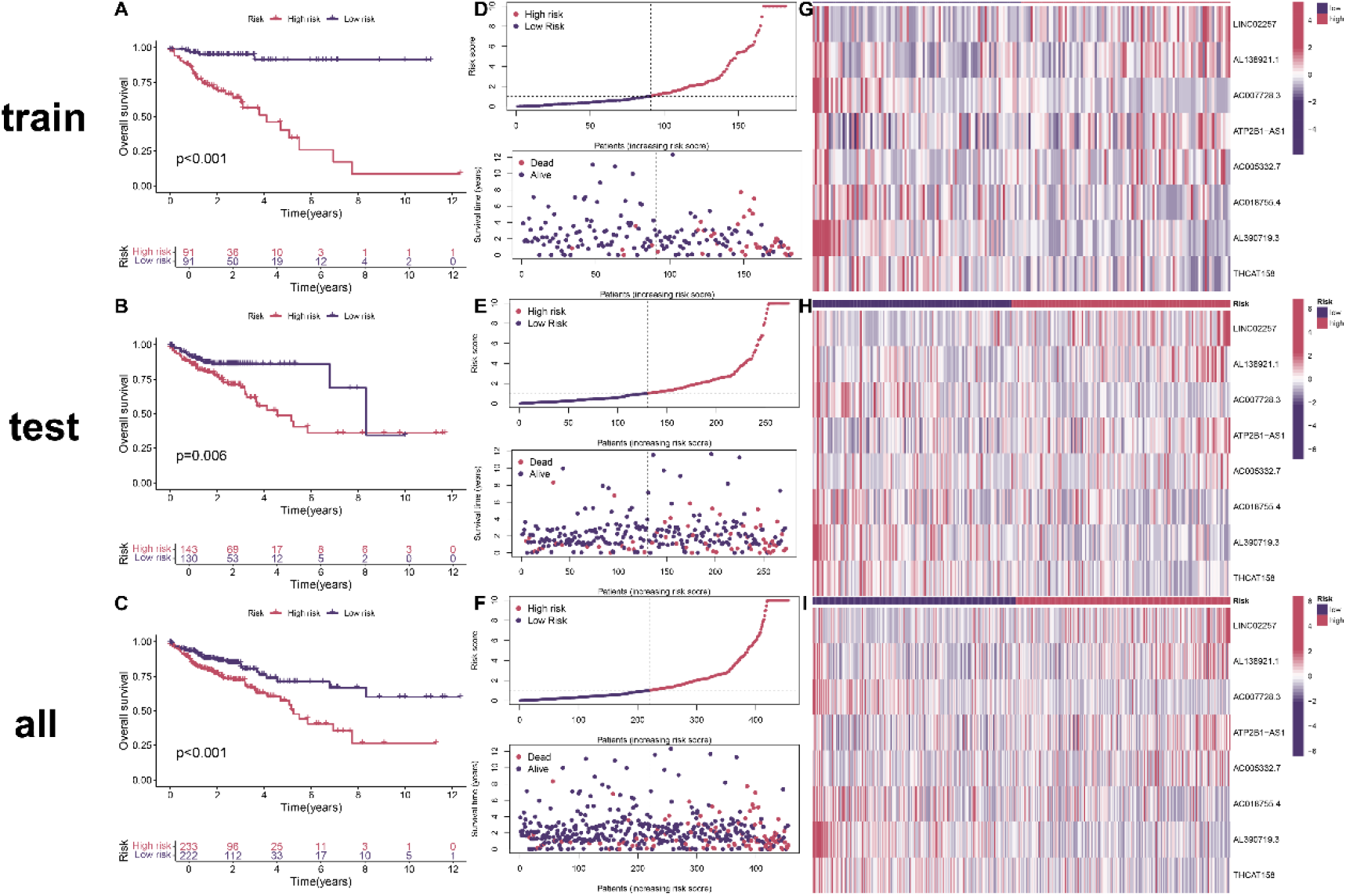
Survival status and risk scores for the total dataset, test set, and training set A-C. Kaplan-Meier survival analysis; D-F. Scatterplots of the distribution of RiskScore values and survival status for colon cancer samples; G-H. Heatmap of differential expression of aging-associated lncRNAs.

**Fig. 4.**
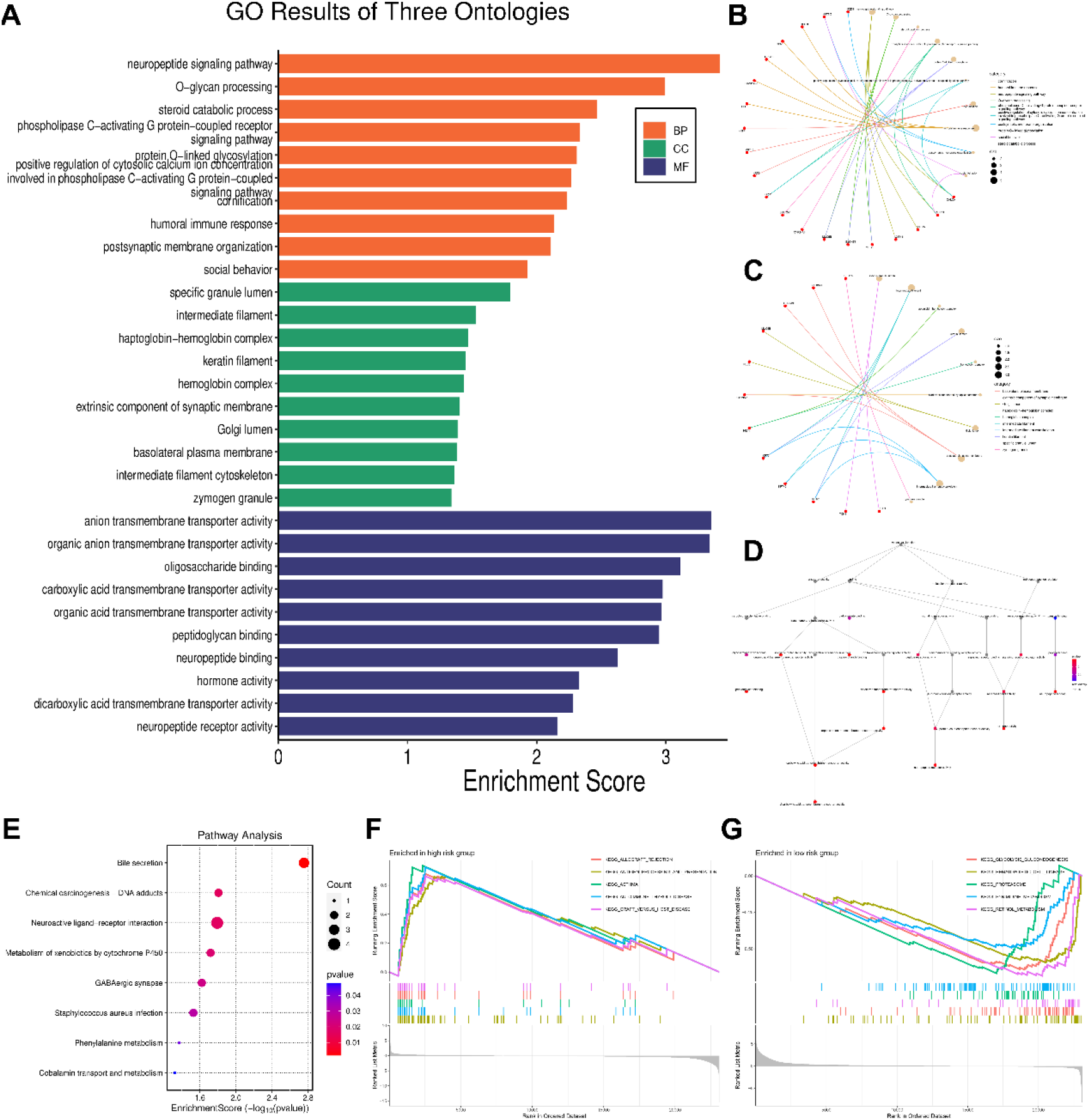
Enrichment analysis A. GO enrichment analysis bar graph; B. Network diagram showing specific enrichment biological process; C. Network diagram showing specific enrichment cellular component; D. Network diagram showing specific enrichment molecular function; E. KEGG pathway enrichment analysis; F. GSEA enriched in high risk group; F. GSEA enriched in low risk group.

### 3.3 Survival analysis of prognostic models

In order to investigate the prognostic evaluation ability of the model, the same method as the training set was used to calculate the risk score for each patient in the test set. Then, based on the median risk score of the training set, all patients in the dataset, training set, and testing set were divided into high-risk and low-risk groups. Survival analysis showed that in the three datasets, patients in the high-risk group had higher mortality rates and shorter overall survival (OS) compared to those in the low-risk group (Fig. 3A-C), indicating that the Senescence-related lncRNA prognostic model has strong predictive ability for colon cancer prognosis. The risk curve shows that the mortality rate of high-risk patients is higher than that of low-risk patients (Fig. 3D-F). The heatmap showed the differential expression of Senescence-related lncRNAs in two groups, with LINC02257, AL138921.1, ATP2B1-AS1, and AC005332.7 being highly expressed in the high-risk group, while AC007728.3, AC018755.4, AL390719.3, and THCAT158 were highly expressed in the high-risk group (Fig. 3G-I).

### 3.4 Immune microenvironment analysis and drug sensitivity analysis

The results of tumor microenvironment analysis showed that the immune score, stromal score, and total score of high-risk OS patients were higher than those of low-risk group patients (Fig. 5A). High risk patients showed higher sensitivity to selective ATP competitive Aurora A and ibrutinib (Fig. 5C, D), which may be potential therapeutic drugs for OS. However, the immune cell infiltration boxplot (Fig. 5E) showed no significant difference in immune cells between high and low-risk groups of patients.

**Fig. 5.**
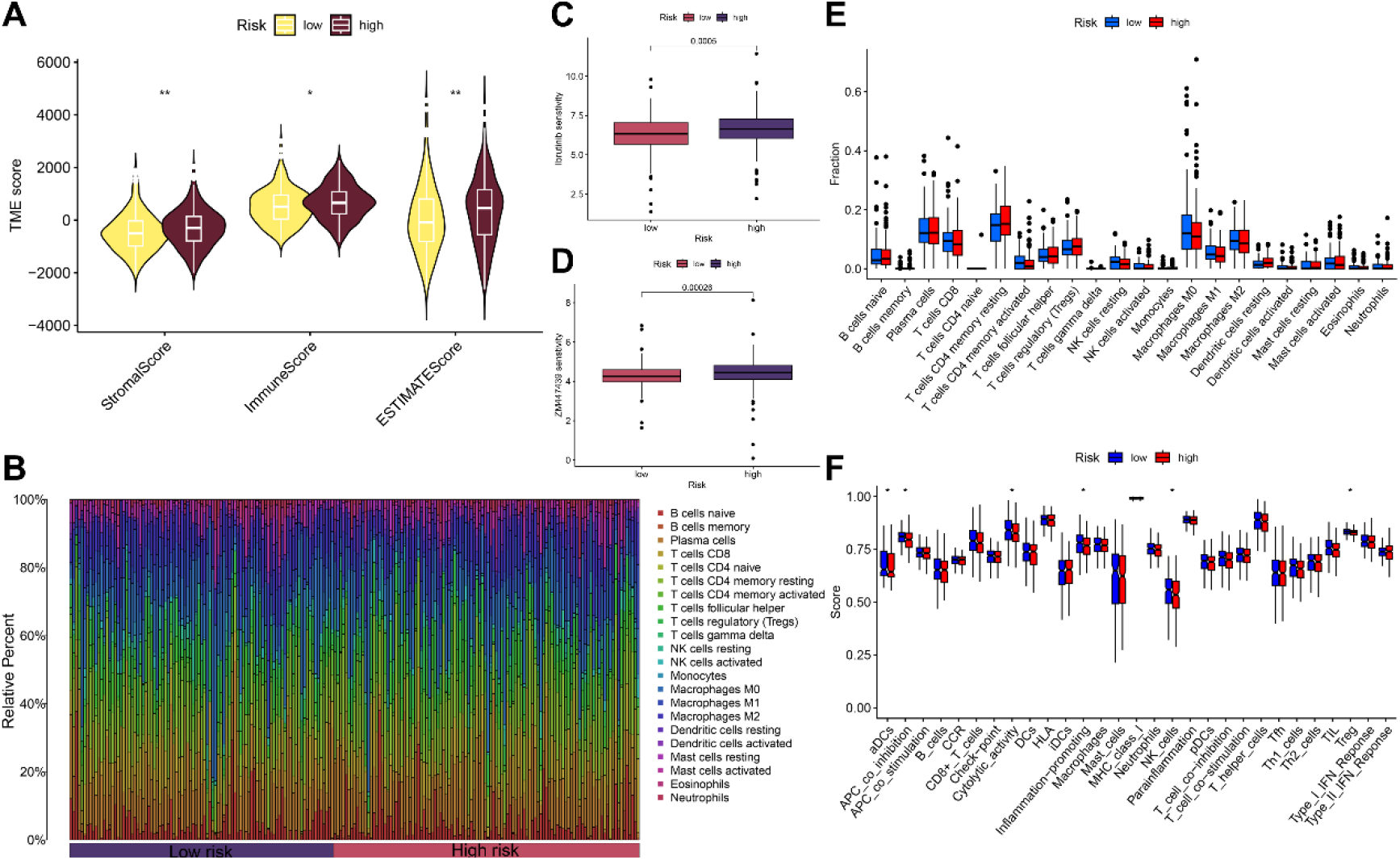
Immune infiltration and drug sensitivity analysis A. Differences in tumor microenvironment between high and low risk groups; C, D. Drug sensitivity analysis between patients in high and low risk groups; E. Results of immune cell infiltration analysis; F. Results of analysis of differences in immune function; *: *P* < 0.05; **: *P* < 0.01; ***: *P* < 0.001

## 4. Discuss

Aging represents a cellular program characterized by the stable growth arrest of damaged or senescent cells. During the growth and development of organisms, processes such as normal embryonic development, tissue remodeling, and wound healing are essential. Tumor cells are also capable of undergoing aging. A key feature of tumor cells is their replication immortality and unchecked proliferation, which suggests that these cells have circumvented the aging processes induced by oncogenes and telomere shortening[10]. A recent study has found that the potential use of anti-aging therapies (known as TIS) for treating colorectal cancer (CRC) requires lower doses compared to inducing cell apoptosis[11].

Previous models for predicting the prognosis of Senescence-related genes in colon cancer have been established using bioinformatics methods[12]. This study established a scoring formula and chemotherapy drug sensitivity model for predicting the prognosis of colon cancer based on Senescence-related LncRNA expression levels using bioinformatics methods. Combined with the LncRNA model, the efficacy of chemotherapy drugs was validated, providing reference for precision medication and prognosis analysis strategies for colon cancer patients.

This study obtained 8 Senescence-related lncRNAs and constructed a prognostic model through co expression analysis, Lasso regression analysis, Cox univariate and multivariate regression analysis. The results of ROC curve, survival analysis, column chart, and heatmap showed that 8 Senescence-related lncRNAs can accurately distinguish the prognosis of high-risk and low-risk groups of patients, and have been validated in early and late stage patients, reliably predicting the outcome of colon cancer patients. In addition, these lncRNAs are prognostic factors independent of other clinical features such as gender and age. Among the 8 Senescence-related lncRNAs in the prognostic model, LINC02257 was previously considered a key lncRNA affecting the prognosis of colon cancer, and can also play a critical role in the proliferation and metastasis of CRC cells through the LINC02257/JNK axis [13-15]. AL138921.1 has played an important role as a key factor in m6A methylation in N staging, survival time, and immune landscape of colon cancer[16]. Previous studies have reported that ATP2B1-AS1 is an important LncRNA associated with disulfide mediated cell death, playing a critical regulatory role in colon cancer, lung adenocarcinoma, and esophageal squamous cell carcinoma[17-19]. It is interesting that AC007728.3 and ATP2B1-AS1 are both LncRNAs involved in disulfide mediated cell death in colon cancer[17]. Unfortunately, there are currently no reports on research on THCAT158, AL390719.3, AC005332.7, and AC018755.4, which will be one of the future research topics.

In summary, this study explored and discovered Senescence-related lncRNA biomarkers that can be used to predict the prognosis of colon cancer. However, there are still many shortcomings in this study. Firstly, the research data comes from the TCGA database, and these retrospective data may have selection bias. Therefore, we also need to validate these results in large multi-center queues. In addition, in order to verify the pathological mechanism of colon cancer discovered and revealed, in-depth research on predicting the efficacy of chemotherapy drugs based on the established model has not been conducted through in vitro cell or model reliability testing. In summary, this study explores the potential interaction between lncRNA and aging to identify potential prognostic markers and search for predictive and therapeutic targets for colon cancer.

## Statements & Declarations

### Author Contributions

All authors contributed to the study conception and design. Material preparation, data collection and analysis were performed by Guangtao Min, Hongpeng Wang. The first draft of the manuscript was written by Yichu Huang and all authors commented on previous versions of the manuscript. All authors read and approved the final manuscript.

### Data Availability

All GWAS summary data that support the findings of this study are openly available in the TCGA and GEO datebases.

